# MultiVirusConsensus: An accurate and efficient open-source pipeline for identification and consensus sequence generation of multiple viruses from mixed samples

**DOI:** 10.64898/2026.03.24.26349218

**Authors:** Niema Moshiri

**Affiliations:** Department of Computer Science & Engineering, UC San Diego, La Jolla, 92093, USA

## Abstract

**Motivation:** Viral surveillance from mixed samples (e.g. wastewater) has become critical in public health efforts to track and contain pathogens. However, existing open-source bioinformatics tools for viral consensus sequence generation are optimized for individual viruses (rather than multiple potential viruses of interest).

**Results:** MultiVirusConsensus is an accurate and efficient open-source pipeline for identification and consensus sequence generation of multiple viruses from mixed samples. It utilizes the memory-efficient ViralConsensus tool via bash process substitution to simultaneously perform consensus sequence calling on all viruses of interest (1) completely in parallel, and (2) by piping datastreams between tools without writing/reading intermediate files (thus eliminating slowdowns related to slow disk accesses).

**Availability:** MultiVirusConsensus is freely available as an open-source software project at: https://github.com/niemasd/MultiVirusConsensus

## 1 Introduction

The epidemiological applications of viral sequencing have expanded rapidly as a result of the COVID-19 pandemic. The real-time reconstruction of viral genomes collected from individual patients (Moshiri, 2023; Ji *et al*., 2024) as well as mixed wastewater samples (Karthikeyan *et al*., 2022) are able to inform real-time public health intervention (Oude Munnink *et al*., 2021; Fielding-Miller *et al*., 2023; Matteson *et al*., 2023; Keehner *et al*., 2024; Wertheim *et al*., 2025). While the majority of recent efforts in this space have focused on a single virus of interest at a time, recent innovations such as the Illumina Viral Surveillance Panel have enabled the simultaneous sequencing of multiple viruses of interest (Shamblin *et al*., 2026).

The reconstruction of consensus genome sequences from raw sequence data requires the use of various bioinformatics pipelines such as iVar (Grubaugh *et al*., 2019), HAVoC (Truong Nguyen *et al*., 2021), V-pipe (Posada-Céspedes *et al*., 2021), ViReflow (Moshiri *et al*., 2022), and many more. However, to our knowledge, all existing open-source viral genome reconstruction pipelines are intended to reconstruct the genome of a *single* virus of interest in a single run, and the only existing pipeline for reconstructing the genomes of *multiple* viruses of interest in a single run is Illumina’s commercial BaseSpace “DRAGEN Microbial Enrichment Plus App” that accompanies their Viral Surveillance Panel.

Here, we introduce MultiVirusConsensus, an accurate and efficient open-source pipeline for identification and consensus sequence generation of multiple viruses from mixed samples. ViralConsensus is fast and memory-efficient and can even be executed with hundreds of viruses of interest on a laptop, making it an ideal tool for real-time viral molecular surveillance.

## 2 Results and discussion

MultiVirusConsensus is a command-line tool written in Python that requires Bash (Ramey, 1994), Minimap2 (Li, 2018), Samtools (Li *et al*., 2009), ViralConsensus (Moshiri, 2023), and optionally BioBloom (Chu *et al*., 2014). MultiVirusConsensus takes the following as required input: (i) one or more FASTQ files containing the reads, (ii) one or more FASTA files containing one or more viral reference genome sequences, and (iii) an output directory to write the results. Optionally, the user can also provide the following: (i) one or more BED files containing amplicon sequencing primers with respect to the provided reference genome sequences for optional primer trimming, (ii) a BioBloom filter (Chu *et al*., 2014) constructed from host (e.g. human) genome sequences for optional host filtering, (iii) the maximum number of threads to use (default: max), (iv) how to handle multimapped reads (“all” to keep all mappings, “best” to keep only the single best mapping, or “none” to discard multimapped reads; default: “all”), and (v) the paths to all dependency executables (default: assume they’re in the user’s `PATH`).

The MultiVirusConsensus pipeline is visualized in Figure 1. First, the reference genomes are loaded from the input FASTA files, and they are merged into a single FASTA file (for use during read mapping) as well as into separate FASTA files with a single reference sequence per file (for use during consensus sequence calling). Next, if provided, the primer BED files are loaded and written into separate BED files with a single reference genome’s primers per file. Then, a Bash script is written to implement the actual pipeline: (i) the reads are mapped to the merged reference genome FASTA using Minimap2, (ii) the read mapping results are written to a BAM file using Samtools, and (iii) the reads are separated into individual consensus sequence calling operations (one per viral reference genome sequence) using ViralConsensus. Importantly, the Bash script itself is included in the output folder, thus ensuring that the analysis is reproducible.

**Fig. 1.**
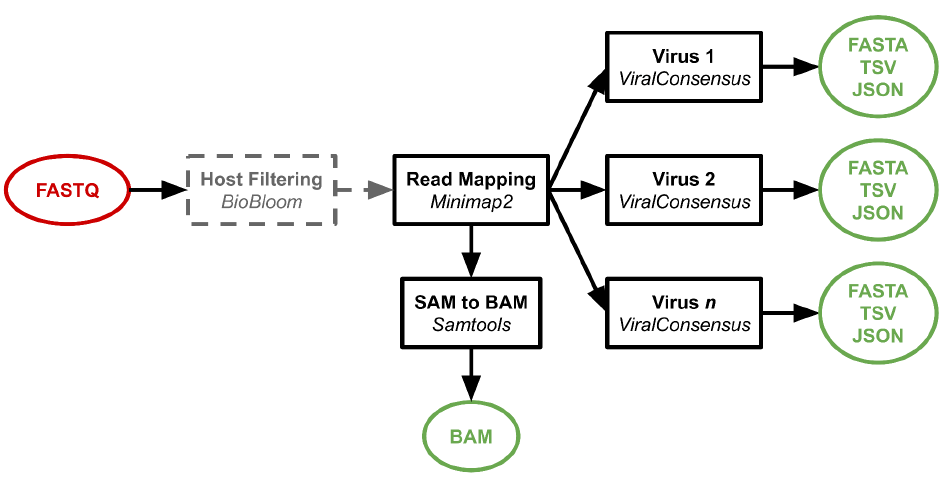
MultiVirusConsensus Pipeline. Diagram depicting the MultiVirusConsensus pipeline. FASTQ files are optionally fed to BioBloom (for host filtering), then to Minimap2 (for read mapping), then simultaneously fed to Samtools (for writing a BAM) as well as to a separate ViralConsensus processes per viral reference genome sequence (for consensus sequence calling).

Importantly, the Bash script produced by MultiVirusConsensus utilizes process substitution in order to avoid writing and subsequently reading intermediate files at each compute-intensive step: the reads are loaded from disk at the beginning of the pipeline, and aside from small reference genome files (KBs per file), all subsequent data movement occurs directly between processes via pipes, until output files are ultimately written to disk at the ends of the pipeline.

In order to assist users as they interpret the results produced by MultiVirusConsensus, we also provide a companion web application: https://niema.net/MultiVirusConsensus (Fig. 2). Users can simply select a MultiVirusConsensus output folder, and the web application will load the results and produce interactive coverage plots that are sorted in descending order of consensus genome completeness (i.e., the number of unambiguous bases in the consensus genome divided by the length of the reference genome). Importantly, the user’s data are not sent anywhere external: the web application is fully client-side (and thus secure), and it can even be executed locally by simply downloading and opening its HTML file. This is important for HIPAA compliance, as users who skip host filtering could accidentally include patient-identifiable information in the BAM file (which includes reads that failed to map to the viral reference genome sequences and thus may contain reads from the host).

**Fig. 2.**
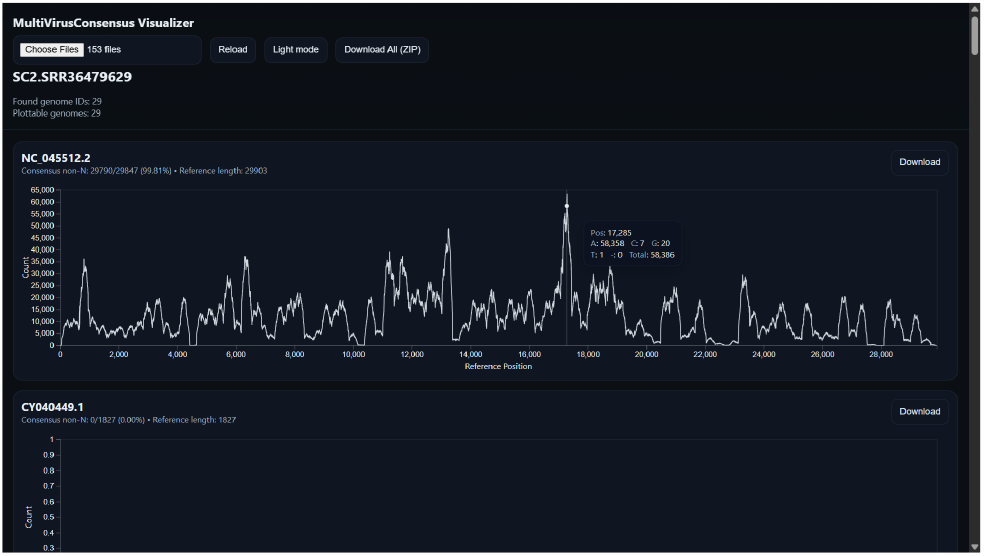
MultiVirusConsensus Companion Web Application. Screenshot demonstrating the MultiVirusConsensus companion web application. The screenshot shows the web application visualizing the results from running MultiVirusConsensus on a real SARS-CoV-2 sequencing dataset: as expected, the top (highest-completeness) consensus genome is SARS-CoV-2, which is the only coverage plot showing significant coverage.

In order to benchmark MultiVirusConsensus, we first built a reference sequence collection consisting of the following 29 sequences: CY040449.1–56.1 (Influenza B Segments 1–8), NC_007366.1–73.1 (Influenza A Segments 1–8), NC_026431.1–38.1 (Influenza A Segments 1–8 Coding Sequences), NC_045512.2 (SARS-CoV-2), OP890336.1 (RSV A), OP965707.1 (RSV B), NC_039199.1 (HMPV), and NC_023891.1 (HPV). We then evaluated the runtime, peak memory, and accuracy of MultiVirusConsensus by processing one simulated and four real sequencing datasets. The simulated dataset was produced from the reference sequence collection by using ART version “MountRainier 2016-06-05” (Huang *et al*., 2012) to simulate Illumina HiSeq 2500 paired-end short reads at 1000X coverage. The real datasets were obtained by selecting four respiratory viruses with similar symptoms that are of typical interest to public health efforts (HMPV, Influenza A, RSV, and SARS-CoV-2) and searching the Sequence Read Archive (SRA) for Illumina MiSeq datasets containing just one of the viruses of interest: SRR8776440 (HMPV), SRR37308007 (Influenza A), SRR37377179 (RSV), and SRR36479629 (SARS-CoV-2).

We then ran MultiVirusConsensus v0.0.5 using its default settings on all four real datasets, a “mixed” real dataset produced by concatenating the four real datasets, and the simulated dataset. For the real datasets, a read mapping was determined to be “correct” if it mapped to the virus already known to be in the sample, or “incorrect” if it mapped to any other reference genome in the collection. For the simulated dataset, a read mapping was determined to be “correct” if it mapped to the reference sequence from which it was simulated, or “incorrect” if it mapped to any other reference sequence in the collection. All datasets and their respective outputs can be found in the following GitHub repository: https://github.com/niemasd/MultiVirusConsensus-Paper

Note that the total number of mapped reads (correct + incorrect) can exceed the total number of reads because reads were allowed to multimap. This is the default behavior in MultiVirusConsensus because homology between related viruses can result in a read mapping well to multiple reference sequences: our intention is to include *all* such mappings, and the user’s downstream coverage analysis (e.g. using the companion web application; Fig. 2) will clarify what viruses are actually likely to exist in the sample. However, this behavior can be overridden via the `--keep_multimapped` command line argument: “all” will keep all good mappings, “best” will keep only the single best mapping, and “none” will discard multimapped reads entirely.

Also, note that the HMPV and SARS-CoV-2 datasets were produced via amplicon sequencing (but were already primer-trimmed), whereas the Influenza A and RSV datasets were not. We expect non-amplicon datasets to have lower proportions of their reads map successfully: accuracy should be assessed by comparing the ratio of the number of correctly-mapped reads vs. the number of incorrectly-mapped reads.

As can be seen in Table 1, across all datasets, the number of reads that map to the correct reference sequence is orders of magnitude larger than the number of reads that map to an incorrect reference sequence, especially in amplicon sequencing datasets. Importantly, this holds even in mixed samples (both real and simulated).

**Table 1.** Benchmark Results. Number of reads, number of correctly mapped reads, number of incorrectly mapped reads, runtime (in seconds), and peak memory usage (in kilobytes) for the four real datasets (HMPV, Influenza A, RSV, and SARS-CoV-2), the “mixed” real dataset, and the simulated dataset. All benchmarks were run on a 2.8 GHz Intel i7-1165G7 CPU with 8 cores and 16 GB of memory.

|  | Reads | Correct | Incorrect | Time (s) | Mem (KB) |
| --- | --- | --- | --- | --- | --- |
| HMPV | 962,668 | 481,943 | 3,582 | 36.37 | 427,128 |
| Flu A | 1,364,614 | 123,430 | 128 | 28.57 | 437,104 |
| RSV | 1,297,542 | 271,163 | 1,196 | 50.88 | 433,452 |
| SC2 | 2,631,924 | 2,629,527 | 0 | 87.54 | 651,380 |
| Mix | 6,256,748 | 3,506,063 | 4,906 | 228.02 | 649,108 |
| Sim | 797,000 | 796,965 | 7,998 | 21.28 | 456,732 |

The runtime, which scales roughly linearly with respect to sequencing dataset size and is roughly constant with respect to reference genome collection size, ranged between 21 seconds and 4 minutes despite extremely high coverage in the datasets.

The peak memory usage, which should scale roughly linearly with respect to reference genome collection size and which should be roughly constant with respect to sequencing dataset size, ranged between 427–652 MB. The fact that there was any variation in the peak memory usage despite using the same reference genome collection is surprising, as the largest memory usage should be the result of having a separate ViralConsensus run for each reference genome running in parallel (and the peak memory usage of ViralConsensus is linear with respect to reference genome length due to the memory allocation of counters for each reference position). The most likely reason for the ~225 MB variation in peak memory usage across the runs, despite using the same reference genome, can likely be attributed to many of the ViralConsensus runs terminating very quickly (potentially before others spawn) when the sequencing dataset size is relatively small (where “small” is actually larger than 1000X coverage). Nevertheless, in all benchmarks, MultiVirusConsensus ran with a peak memory usage far lower than 1 GB, meaning this 29-reference collection can be comfortably executed on lightweight devices (e.g. a Raspberry Pi).

In sum, we introduce MultiVirusConsensus, an accurate and efficient open-source pipeline for identification and consensus sequence generation of multiple viruses from mixed samples. ViralConsensus is fast and memory-efficient and can even be executed with hundreds of viruses of interest on a laptop, making it an ideal tool for real-time viral molecular surveillance. We hope ViralConsensus will aid viral molecular epidemiologists in their efforts to analyze viral sequence data at massive scale, particularly as an open-source alternative to similar commercial pipelines such as Illumina’s BaseSpace “DRAGEN Microbial Enrichment Plus App” that accompanies their Viral Surveillance Panel.

## Data Availability

All data produced are available online at: https://github.com/niemasd/MultiVirusConsensus-Paper

https://github.com/niemasd/MultiVirusConsensus

https://github.com/niemasd/MultiVirusConsensus-Paper

## Acknowledgements

We would like to thank Louise Laurent, Peter De Hoff, and Rob Knight for fruitful conversations.

## References

Chu, J. et al. (2014) BioBloom tools: fast, accurate and memory-efficient host species sequence screening using bloom filters. Bioinformatics, 30(23), 3402–3404.

Fielding-Miller, R.K. et al. (2023) Safer at School Early Alert: An observational study of wastewater and surface monitoring to detect COVID-19 in elementary schools. The Lancet Regional Health - Americas, 19, 100449.

Huang, W. et al. (2012) ART: a next-generation sequencing read simulator. Bioinformatics, 28(4), 593–594.

Ji, D. et al. (2024) ViralWasm: a client-side user-friendly web application suite for viral genomics. Bioinformatics, 40(1), btae018.

Karthikeyan, S. et al. (2022) Wastewater sequencing reveals early cryptic SARS-CoV-2 variant transmission. Nature, 609, 101–108.

Keehner, J. et al. (2024) Integrated Genomic and Social Network Analyses of Severe Acute Respiratory Syndrome Coronavirus 2 Transmission in the Healthcare Setting. Clinical Infectious Diseases, 78(5):1204–1213.

Li, H. (2018) Minimap2: pairwise alignment for nucleotide sequences. Bioinformatics, 34(18), 3094–3100.

Li, H. et al. (2009) The Sequence Alignment/Map format and SAMtools. Bioinformatics, 25(16), 2078–2079.

Matteson, N.L. et al. (2023) Genomic surveillance reveals dynamic shifts in the connectivity of COVID-19 epidemics. Cell, 186(26):5690–5704.e20.

Moshiri, N. (2023) ViralConsensus: A fast and memory-efficient tool for calling viral consensus genome sequences directly from read alignment data. Bioinformatics, 39(5), btad317.

Moshiri, N. et al. (2022) The ViReflow pipeline enables user friendly large scale viral consensus genome reconstruction. Sci. Rep., 12, 5077.

Oude Munnink, B.B. et al. (2021) The next phase of SARS-CoV-2 surveillance: real-time molecular epidemiology. Nat. Med., 27, 1518–1524.

Posada-Céspedes, S. et al. (2021) V-pipe: a computational pipeline for assessing viral genetic diversity from high-throughput data. Bioinformatics, 37(12), 1673–1680.

Ramey, C. (1994) Bash, the Bourne-Again Shell. Free Software Foundation.

Shamblin, A. et al. (2026) Simultaneous detection of multiple viral pathogens in vacuumed dust from different building types. Building and Environment, 114530,

Truong Nguyen, P.T. et al. (2021) HAVoC, a bioinformatic pipeline for reference-based consensus assembly and lineage assignment for SARS-CoV-2 sequences. BMC Bioinf., 22, 373.

Wertheim, J.O. et al. (2025) Phylogeographic and genetic network assessment of COVID-19 mitigation protocols on SARS-CoV-2 transmission in university campus residences. eBioMedicine, 116:105729.

Yang, C. et al. (2017) NanoSim: nanopore sequence read simulator based on statistical characterization. GigaScience, 6(4), gix010.

